# Preclinical Trial Results of Main Pancreatic Duct Endoluminal Radiofrequency Ablation to Reduce Postoperative Pancreatic Fistula

**DOI:** 10.64898/2026.05.01.26352130

**Authors:** Gemma Vellalta, Francesca Marcucci, Patricia Sánchez-Velázquez, Enrique Berjano, Anna Andaluz, Fernando Burdio, Benedetto Ielpo

**Author notes:** **Corresponding author:** Benedetto Ielpo, Hepato-Biliary and Pancreatic Surgery Unit, Department of Surgery, Hospital Parc Salut Mar, Barcelona, Spain. Address: Passeig Maritim 25, 08034, Barcelona, Spain. **Funding:** FIS PI23/00445 / Health Institute Carlos III (Spanish Ministry of Economy, Industry and Innovation) (FIS Grant) Grant PID2022-136273OB-C32 (Spanish Ministerio de Ciencia e Innovación, Agencia Estatal de Investigación, co-funded by the European Regional Development Fund). **Previous communication to a society or meeting** This study has not been previously published and is not under consideration elsewhere. Parts of this work may derive from experimental research previously presented in preliminary form within the context of institutional or academic meetings, but no abstract or full communication containing the data presented here has been formally published. **Data availability statement** The data supporting the findings of this study are available from the corresponding author upon reasonable request. The data are not publicly available due to their nature as preclinical experimental data generated in a controlled animal research setting.

## Abstract

**Background:** Postoperative pancreatic fistula (POPF) is a major cause of morbidity after pancreatoduodenectomy, particularly in patients with high-risk pancreatic remnants. Preventive strategies based solely on surgical technique have yielded inconsistent results, and thus there has been growing interest in strategies aiming to modify the biological behavior of the pancreatic remnant. This preclinical study evaluated the biological and histopathological effects of preoperative endoluminal radiofrequency ablation (ERFA) of the main pancreatic duct (MPD) performed 4 weeks before pancreatic transection in a porcine model.

**Methods:** Animals underwent laparoscopic MPD occlusion followed by pancreatic transection at 4 weeks and necropsy 15 days thereafter. Feasibility, safety, histological atrophy, and macroscopic findings associated with POPF risk were assessed. As a secondary objective, outcomes were compared with a that underwent MPD occlusion using cyanoacrylate glue.

**Results:** Preoperative ERFA was technically feasible and safe. At 4 weeks, ERFA induced marked and homogeneous acinar atrophy that was significantly greater than that observed after glue occlusion (p = 0.018), indicating effective biological conditioning of the pancreatic remnant. At necropsy, pseudocyst formation and intra-abdominal adhesions, known surrogate markers of pancreatic fistula in pigs, were significantly more frequent in the glue group and absent in ERFA-treated animals. Serum amylase levels, postoperative weight gain, complication rates, and preservation of endocrine architecture were comparable between groups.

**Conclusions:** Ductal ablation of the MPD via ERFA induced stable, progressive exocrine pancreatic atrophy, effectively preconditioning the gland prior to pancreatic transection. Experimental evidence suggests that its biological effects stabilize approximately 4 weeks after treatment. Compared to cyanoacrylate occlusion, ERFA achieved more homogeneous early biological effects and fewer fistula-related macroscopic complications. These findings support the further investigation of preoperative pancreatic conditioning as a potential adjunct strategy for POPF risk reduction, although clinical studies are needed to clarify its role alongside established reconstructive approaches.

## Introduction

Postoperative pancreatic fistula (POPF) remains the principal determinant of morbidity after pancreatoduodenectomy (PD) despite substantial advances in surgical technique, perioperative care, and centralization of pancreatic surgery ^1^. Although perioperative mortality has fallen below 5% in high-volume centers, up to 20%–30% of patients still develop clinically relevant POPF, particularly in the presence of a soft pancreatic remnant and a small main pancreatic duct ^2-4^. It should be noted that POPF is not merely a local complication but rather a trigger for downstream events including hemorrhage, intra-abdominal sepsis, delayed gastric emptying, prolonged hospital stay, and delayed or omitted adjuvant therapy in oncological patients.

Efforts to reduce POPF have traditionally focused on the refinement of pancreatico-enteric anastomosis. However, none of the anastomotic techniques, such as pancreaticojejunostomy or pancreaticogastrostomy with or without stents, have consistently demonstrated superiority in preventing clinically relevant POPF ^5,6,7^. The persistent failure of these purely technical solutions has sparked interest in alternative strategies aimed at modifying the biological behavior of the pancreatic remnant itself.

One such strategy is pancreatic exclusion by occluding the main pancreatic duct (MPD), thereby bypassing the need for fragile anastomosis. MPD occlusion has been explored for several decades, initially through duct ligation and later through the intraductal injection of sealing agents (i.e., synthetic or biological glues). In theory, this should suppress exocrine secretion, induce acinar atrophy, and thereby reduce enzymatic injury at the pancreatic stump while preserving endocrine function. However, despite this appealing concept, clinical results have been inconsistent ^8-12^. Some problems of duct ligation include incomplete sealing and severe fistula formation ^8^, while glue occlusion (GO) has shown variable rates of duct recanalization and, in some series, long-term endocrine impairment ^8,9^. Consequently, pancreatic exclusion is not yet widely accepted and remains reserved for selected high-risk scenarios.

Given this context, energy-based approaches have emerged as promising alternatives. Similar to glue injection, these methods target the biological mechanisms underlying POPF rather than aiming for mechanical reconstruction alone ^13-17^. One such method is endoductal radiofrequency ablation (ERFA), which uses controlled thermal energy to induce ductal epithelial necrosis, fibrosis, and durable closure, leading to progressive exocrine atrophy while preserving endocrine islet function^18^.

Our previous experimental porcine studies investigated ERFA of the MPD immediately after pancreatic transection as an alternative strategy for ductal occlusion, and it demonstrated more consistent and durable ductal ablation than glue-based occlusion ^19,20^. We have also reported the successful use of ERFA in selected high-risk patients undergoing PD, wherein no cases of clinically relevant grade C POPF or procedure-related mortalities were observed ^20,21^. Notably, our experimental results revealed that the maximal and biologically stable effect of ductal ablation occurs not immediately, but rather approximately 30 days after treatment, specifically once postablation inflammation has resolved and exocrine atrophy is fully established.

This observation led to a critical conceptual shift. Rather than occluding the duct at the time of pancreatic resection, a preoperative interval may be necessary for the optimal biological conditioning of the pancreas. Preoperative MPD ablation can transform the pancreas from a high-risk, enzyme-secreting gland into a low-secretory, fibrotic remnant at the time of transection, thereby reducing the likelihood and severity of POPF.

This preclinical study was designed to evaluate the safety and biological efficacy of preoperative MPD occlusion via ERFA performed 30 days before pancreatic transection ^22^. The primary objective was to assess whether ERFA induces stable and favorable pancreatic remodeling prior to resection. As a secondary objective, outcomes were compared with those obtained using cyanoacrylate GO, the most common alternative occlusion technique to date. This study provides experimental evidence that supports preoperative pancreatic conditioning as a novel strategy for preventing POPF.

## Methods

This randomized preclinical study involved the use of healthy Landrace pigs (3-4 months old, 30-40 kg) at the CMCiB center, a facility accredited for Good Laboratory Practice surgical studies. The study protocol is based on the original protocol previously published by our group ^22^ and has been approved by the Animal Ethics Committee of the Government of Catalonia (Ref. 22-017-BL).

Animals were randomly assigned to undergo either MPD occlusion via ERFA or GO prior to pancreatic transection. No predefined exclusion criteria were applied. All animals that underwent randomization were included in the analysis, and none were excluded from outcome assessment.

The intervention followed a three-stage approach (Figure_1). First, MPD occlusion was done via hybrid laparoscopic surgery on day 0, using either a 3 Fr radiofrequency ablation catheter (10 W, retrograde application) or 1 ml of N-Butyl-2-Cyanoacrylate plus methylacrylosulfalane (Glubran 2). Second, laparoscopic pancreatic transection was done after 4 weeks, taking a sample of the level of transection for histopathological analysis. Third, necropsy with histopathological analysis was done 15 days posttransection.

**Figure_1:**
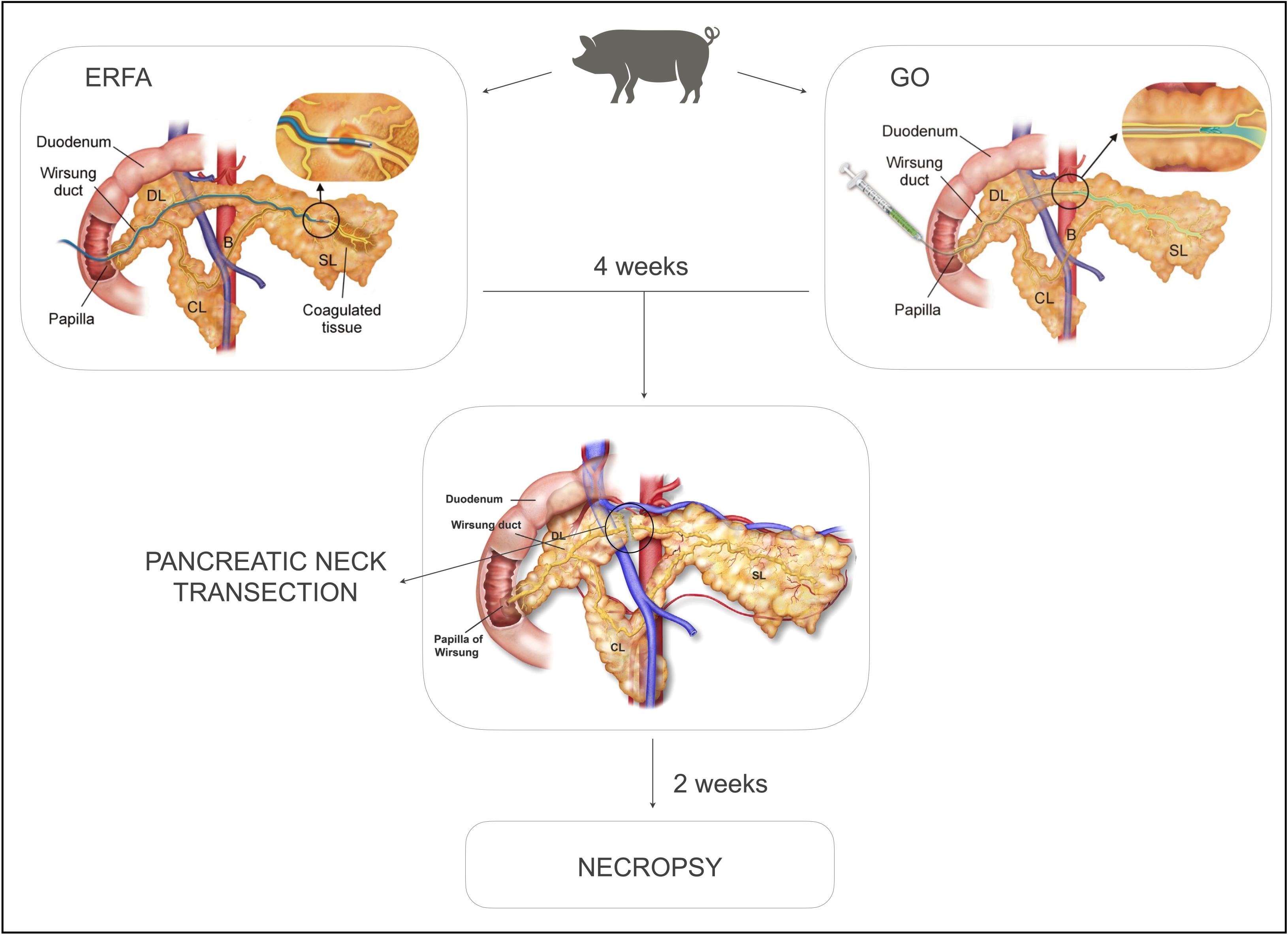
Three-stage approach of main pancreatic duct occlusion followed by pancreatic transection and necropsy.

Regarding clinical monitoring, the animals underwent daily welfare assessments, weekly weight measurements. Serum amylase levels were measured at 48 hours after MPD occlusion, and drainage amylase was measured at 48 hours after pancreatic transection. The animals were euthanized at 15 days posttransection. A midline laparotomy was performed to assess the peritoneal cavity, pancreatic stumps, adhesions, and pseudocysts. The pancreas was excised, and the MPD was cannulated when possible. Specimens were fixed in 10% buffered formalin, processed, and embedded in paraffin.

Histopathological analyses were performed with the support of experienced pathologists who were blinded to treatment allocation. Evaluation included serial sectioning (3 µm) with hematoxylin and eosin staining of the 1) transection specimens and 2) proximal, distal, and accessory pancreatic tissue of the necropsy. As described by Yagi et al., the acinar atrophy were classified according to the percentage of normal pancreatic parenchyma as either grade 0 (90%–100%), grade 1 (70%–89%), grade 2 (30%–69%), or grade 3 (< 29%) ^23^. The presence of Islets of Langerhans was also determined as a marker of endocrine preservation.

Sample size was estimated *a priori* based on expected differences in fistula-related outcomes between groups using the Granmo® calculator (24), estimating 21 animals per group for a clinically relevant difference in POPF rates (50% in GO vs. 11% in ERFA), assuming a two-sided alpha of 0.05 and 80% power. However, due to clear differences in outcomes, the study was stopped early after approximately half of the planned sample, in accordance with ethical principles of animal reduction. In accordance with the early stopping criteria and our ethical and animal welfare guidelines, as well as due to the encouraging preliminary results, we decided to complete the study after reaching 18 pigs (10 in ERFA and 8 in GO) to minimize unnecessary animal use.

Data collection and analysis were conducted without knowledge of group assignment whenever feasible. Statistical analysis consisted of two-sided unidimensional statistical tests, with p <0.05 considered significant. Continuous and categorical variables were analyzed using the Mann–Whitney U test and Fisher’s Exact Test, respectively. Due to the exploratory nature of the study, no adjustments were made for baseline covariates or multiple comparisons. One subject died prematurely from severe sepsis secondary to a colonic lesion caused by the laparoscopic trocar during the second surgery. As this complication was unrelated to the occlusion treatment, the subject (BL03) was excluded from the analysis from the surgery onwards. Additionally, as part of the experimental setup phase, two subjects underwent a robotic ERFA approach to assess feasibility, as prespecified in the initial protocol ^22^. These two preliminary cases were not included in the randomized study since a different surgical technique was used.

## Results

For clarity and brevity, ERFA and GO data are reported side-by-side within each outcome measure. Baseline characteristics and main outcomes are summarized in Table 1.

**Table 1:** Baseline Characteristics and Main Outcomes.

|  | ERFA | GO | *p-value* |
| --- | --- | --- | --- |
| Age (months), mean (SD) | 3.10 (3.00–3.38) | 3.38 (3.00–3.62) | 0.23 |
| Weight (Kg), mean (SD) | 35.70 (32.88–39.25) | 38.95 (36.60–39.25) | 0.48 |
| Weight gain (kg), mean (SD) | 4.00 (1.50–5.50) | 4.62 (1.25–9.25) | 1 |
| Serum Amylase 48h after DO (U.L), mean (SD) | 6701.60 (1102.75–4450.75) | 10701.88 (8284.50–14538.50) | 0.10 |
| Atrophy grade 4 weeks after DO (*), n (%) |  |  | **0.018** |
| 0 | 1 (10.0%) | 4 (50.0%) |  |
| 1 | 1 (10.0%) | 2 (25.0%) |  |
| 2 | 6 (60.0%) | 1 (12.5%) |  |
| 3 | 1 (10.0%) | 0 (0%) |  |
| NA | 1 (10.0%) | 1 (12.5%) |  |
| Distal pancreas atrophy score at necropsy (*), n (%) |  |  | 1 |
| 0 | 1 (11.1%) | 2 (25.0%) |  |
| 1 | 5 (55.6%) | 3 (37.5%) |  |
| 2 | 1 (11.1%) | 1 (12.5%) |  |
| 3 | 0 (0%) | 1 (12.5%) |  |
| NA | 2 (22.2%) | 1 (12.5%) |  |
| Proximal pancreas atrophy score at necropsy (*), n (%) |  |  | 0.72 |
| 0 | 0 (0%) | 3 (37.5%) |  |
| 1 | 2 (22.2%) | 1 (12.5%) |  |
| 2 | 4 (44.4%) | 1 (12.5%) |  |
| 3 | 1 (11.1%) | 3 (37.5%) |  |
| NA | 2 (22.2%) | 0 (0%) |  |
| Minor pancreas atrophy score at necropsy (*), n (%) |  |  | 0.71 |
| 0 | 2 (22.2%) | 3 (37.5%) |  |
| 1 | 1 (11.1%) | 1 (12.5%) |  |
| 2 | 5 (55.6%) | 2 (25.0%) |  |
| 3 | 0 (0%) | 1 (12.5%) |  |
| NA | 1 (11.1%) | 1 (12.5%) |  |
| Pseudocyst, n (%) |  |  | **0.015** |
| 0 | 8 (80.0%) | 2 (25.0%) |  |
| 1 | 1 (10.0%) | 6 (75.0%) |  |
| Adherences, n (%) |  |  | **<0.001** |
| 0 | 9 (90.0%) | 1 (12.5%) |  |
| 1 | 0 (0%) | 7 (87.5%) |  |
| Clavien Dindo, 1st surgery, n (%) |  |  | 0.76 |
| 0 | 6 (60.0%) | 5 (62.5%) |  |
| 1 | 1 (10.0%) | 2 (25.0%) |  |
| 2 | 3 (30.0%) | 1 (12.5%) |  |
| Clavien Dindo, 2nd surgery, n (%) |  |  | 0.27 |
| 0 | 6 (60.0%) | 3 (37.5%) |  |
| 1 | 2 (20.0%) | 3 (37.5%) |  |
| 2 | 1 (10.0%) | 2 (25.0%) |  |

(\*) Acinar atrophy according to the parenchymal atrophy classification of Yagi et al 2018: percentage of normal pancreatic parenchyma grade 0 (90%–100%), grade 1 (70%–89%), grade 2 (30%–69%), or grade 3 (< 29%). SD: Standard Deviation. NA: Not Available.

### Baseline Characteristics

The study population was well-balanced at baseline. There were no significant differences between groups in terms of sex distribution (Fisher’s exact test, p = 1.000; OR = 1.47, 95% CI: 0.16–14.14) and age (GO: 3.38 ± SD years vs. ERFA: 3.10 ± SD years). Non-parametric (Wilcoxon rank–sum, p = 0.231) and parametric (Welch’s t-test, p = 0.217) analyses confirmed the absence of significant differences. Baseline body weight was also comparable (GO: 38.95 kg vs. ERFA: 35.70 kg), with no significant differences on Wilcoxon test (p = 0.475) or t-test (p = 0.195). Scatterplots of weight versus age demonstrated similar trends in both groups, with no evidence of group-specific interactions. Thus, the demographic and physiological characteristics were well matched prior to intervention.

### Treatment Effects on Pancreatic Atrophy

Early histological evaluation at 4 weeks after MPD occlusion revealed a significant difference in acinar atrophy between the groups. Using a 0–3 ordinal scale (Yagi et al., 2018) and the Mann–Whitney U test, animals in the ERFA group exhibited higher atrophy scores than the GO group (p = 0.018), indicating more extensive acinar tissue loss (Figure 2). This difference was confirmed on violin and boxplot visualizations (Figure 3), wherein the ERFA group demonstrated nearly complete atrophy (score 3) whereas the GO group retained largely intact acinar architecture (score 0).

**Figure 2:**
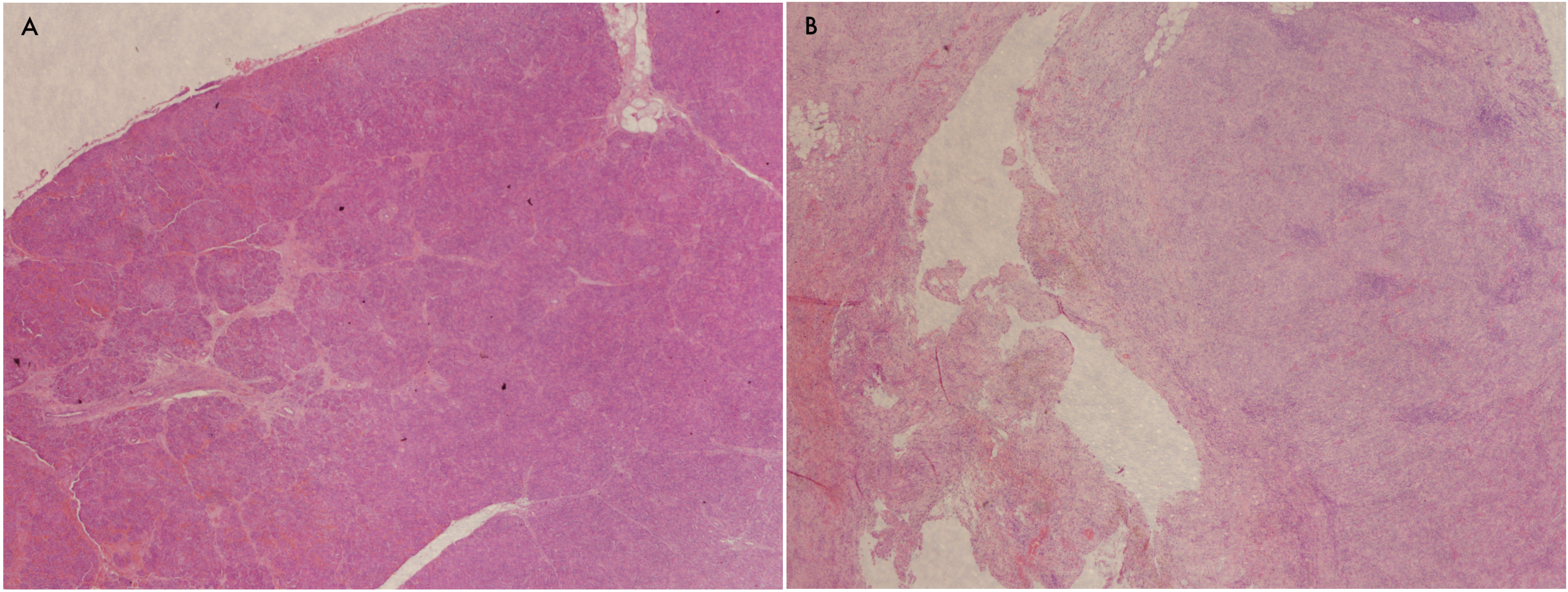
Microscopic pattern of the pancreatic transection sample at 4 weeks. Representative histological features are shown. In these examples, there is a marked loss of the acinar component in the ERFA group, compared to an almost complete preservation of the gland in the GO group. A: Glue group; B: ERFA group.

**Figure 3:**
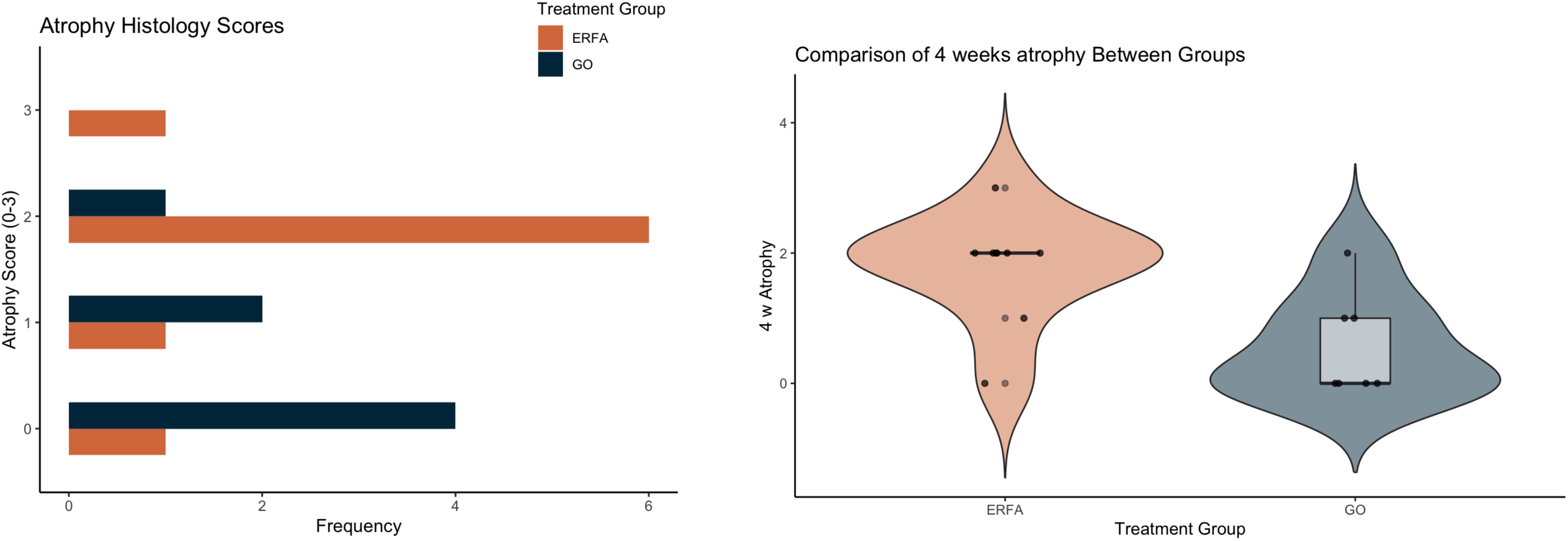
Bar plot comparing pancreatic atrophy scores in endoductal radiofrequency ablation (EFRA) versus glue occlusion (GO).

At necropsy, atrophy was assessed in the proximal, distal, and uncinate regions of the pancreas. No statistically significant differences were observed between the ERFA and GO groups at any anatomical level (proximal: p = 0.474; distal: p = 1.000; uncinate: p = 0.761). However, postoperative tissue remodeling, inflammatory responses, and surgical trauma likely contributed to moderate atrophy in most specimens, potentially masking subtle differences between groups.

### Macroscopic Necropsy Findings: Risk of Fistula

Pseudocysts were significantly more common in the GO group versus the ERFA group (Fisher’s Exact Test, p = 0.015; OR = 0.054, 95% CI: 0.0008–0.787), highlighting the protective effect of ERFA against cyst formation. Notably, unlike in humans, pancreatic fistulas in pigs predominantly manifest through pseudocyst formation. Representative images demonstrated large pseudocysts in the transection area of the GO group, whereas none were observed in the ERFA group.

Adhesion formation was also markedly different. Postoperative adhesions were seen in 7 out of 8 Glue-treated animals, but none were observed in the ERFA group (Fisher’s exact test, p = 0.00041; OR = 0). Pseudocyst formation was strongly associated with the formation of adhesions (Fisher’s exact test, p = 0.0037; phi = 0.636), suggesting that these complications share an underlying pathophysiological mechanism (Figure 4).

**Figure 4:**
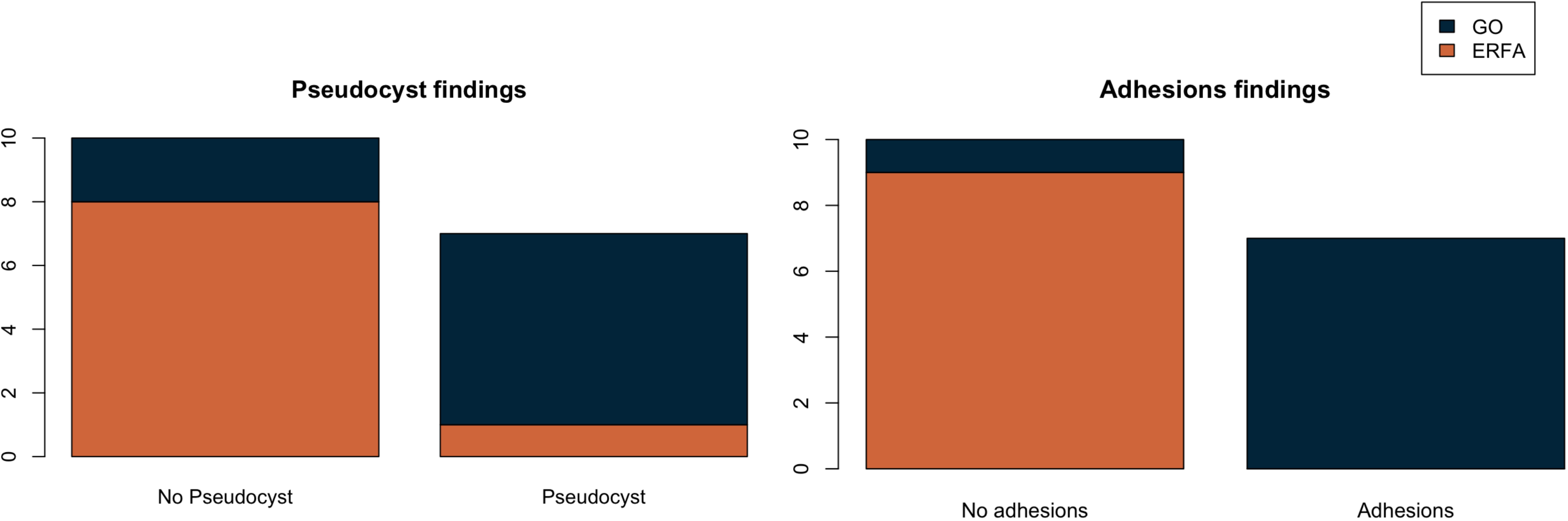
Bar plots comparing the presence of pseudocysts and adhesions in endoductal radiofrequency ablation (EFRA) versus glue occlusion (GO).

### Biochemical and clinical outcomes

Serum amylase measured at 48 hours after duct occlusion did not significantly differ between GO and ERFA groups (W = 59, p = 0.101). Post-treatment weight gain was equivalent between groups (p = 1), reflecting similar nutritional and metabolic recovery.

Quantitative assessment of perioperative complications with the Clavien–Dindo (CD) grading indicated no significant differences between groups. No major perioperative complications were reported, although a small number of wound infections were present, mostly related to animal bites (grade 2 or below). After the first surgical procedure, CD scores were comparable (p = 0.760). Following the second surgery, similar complication profiles were obtained (p = 0.2655).

Collectively, these results indicate well-balanced baseline characteristics, thereby validating treatment comparisons. ERFA induced significantly greater early histological atrophy at 4 weeks. Additionally, ERFA was also associated with markedly lower rates of pseudocyst formation and adhesion development at necropsy, and these complications demonstrated a strong positive association. Late-stage atrophy, serum and drain amylase levels, weight gain, and Clavien–Dindo complication scores did not differ between EFRA and GO, indicating their comparable safety profiles despite differences in early histological and macroscopic outcomes.

## Discussion

POPF remains a major cause of morbidity after PD despite sustained technical refinement. The biological properties of the pancreatic remnant, particularly gland texture and duct size, are key outcome determinants, but preventive efforts have largely focused on anastomotic technique ^25-27^. Accordingly, MPD occlusion has been proposed to suppress exocrine secretion and reduce enzymatic injury, but conventional approaches using duct ligation or intraductal glue have yielded inconsistent results and raised concerns regarding durability and endocrine function.

This preclinical study investigated MPD occlusion via ERFA performed four weeks prior to pancreatic transection. This approach was demonstrated to be technically feasible and safe. It induced a controlled degree of acinar atrophy that is biologically favorable and can sufficiently reduce the risk of downstream complications associated with POPF. At 4 weeks, ERFA caused the early and homogeneous suppression of exocrine activity, resulting in a functionally low-secretory pancreas upon transection. Notably, this is not a novel concept; experimental and clinical studies have previously demonstrated that acute and complete pancreatic duct occlusion does not induce acute pancreatitis per se. Instead, inflammatory complications are more commonly associated with partial or unstable obstruction ^28,29^. This is because durable ductal occlusion activates apoptotic pathways, causing progressive exocrine involution rather than necroinflammatory injury ^15^. This mechanism may explain the higher incidence of pseudocyst formation observed after GO compared with ERFA, since complete ductal sealing is more technically challenging and carries a higher risk of repermeabilization. In contrast, ERFA provides a more homogeneous and easily reproducible seal, thereby reducing enzymatic load at the pancreatic stump and acting through modulation of the pancreatic substrate instead of aiming for anatomical closure through a surgical technique.

Compared to preoperative ERFA, cyanoacrylate GO was effective in partially reducing pancreatic secretion but was less consistent in inducing atrophy at the early four-week timepoint. This could be because mechanical obstruction with cyanoacrylate relies on complete luminal filling and is susceptible to extrusion or micro-recanalization, often resulting in the preservation of acinar architecture and continued enzymatic activity. Consequently, glue has a more variable biological effect, and its capacity to precondition the gland before surgery appears inferior to that of ERFA.

Macroscopic findings at necropsy further support the superiority of ERFA. Pseudocyst formation and intraabdominal adhesions were markedly reduced in the ERFA group versus the GO group. These complications in the GO group likely reflect ongoing leakage of pancreatic juice due to incomplete ductal obstruction, which promote local inflammation, fibro-cystic changes, and adhesion formation. On the other hand, the near-complete absence of these adverse events in the ERFA group underscores the durability and functional efficacy of energy-based ductal ablation. By inducing controlled atrophy and ductal closure before surgery, ERFA not only modifies the tissue substrate to reduce enzymatic exposure but also minimizes local inflammatory sequelae that can complicate postoperative recovery.

Interestingly, although the EFRA group had significantly more pronounced early acinar atrophy, there were less marked differences in the degree of atrophy across pancreatic regions observed at necropsy. This finding suggests that MPD occlusion, whether achieved via ERFA or glue, ultimately leads to exocrine atrophy over time. In the GO group, posttransection tissue remodeling and delayed inflammatory changes likely contributed to progressive obstruction of the remnant, resulting in secondary postoperative atrophy. Nevertheless, these histological assessments were performed only at later time points, which can potentially underestimate differences in the early biological effects of ductal occlusion and may be confounded by fistula-related inflammation.

Although ductal occlusion can induce pancreatic atrophy, our study demonstrated that this process requires time and that its preventive potential depends critically on timing. Ductal occlusion was performed 30 days before pancreatic transection, which allowed sufficient time for controlled exocrine involution to occur, thereby conditioning the gland before surgery. Conversely, performing occlusion during the same operation as resection is too late, because the biological processes leading to fistula formation have already been initiated. In such cases, atrophy cannot yet exert its protective effect, thus poorly mitigating the risk of POPF ^19, 20, 21^.

Another interesting observation in this study is that endocrine function was preserved based on the maintained islet architecture and stable weight gain in both groups. This suggests that ERFA selectively targets exocrine tissue while sparing endocrine cells. However, postoperative diabetes was not directly measured in the porcine model, because this would have required long-term follow-up. Nevertheless, our previous clinical study in human patients ^21^ found no differences in endocrine insufficiency between groups, supporting the translational relevance of endocrine preservation. More human studies are needed to evaluate long-term function preservation.

Regarding safety, pancreatic transection was feasible in all animals after MPD occlusion (both via EFRA and glue) without increased operative difficulty or complications, as quantitatively supported by the consistently low Clavien–Dindo grading. This confirms that preoperative tissue remodeling does not compromise surgical safety or technical feasibility.

A potential future clinical application based on these findings could involve preoperative endoscopic ERFA of the MPD via ERCP approximately 30 days before PD in patients with a high risk of fistula formation. By preoperatively inducing controlled exocrine suppression, this strategy may biologically condition the pancreatic remnant at the time of transection. However, it should be noted that the present study was not designed to compare ductal ablation with pancreato-enteric anastomosis, which remains the current clinical standard. Nevertheless, these results suggest that biological modulation of the remnant may represent a complementary approach to conventional reconstruction. This can be particularly useful in high-risk patients with soft glands, small ducts, or a fatty pancreas, settings wherein conventional strategies often fail, and in frail patients otherwise considered for total pancreatectomy and its associated morbidity ^26-30^.

Previous studies on pancreatic duct ligation and GO have raised concerns regarding endocrine impairment and postoperative diabetes, thereby limiting the widespread adoption of ductal exclusion strategies. However, the preservation of islet architecture and stable postoperative weight gain in this study did not suggest any relevant endocrine dysfunction, consistent with our preliminary clinical experience in high-risk patients. Nevertheless, these observations remain indirect and limited by short follow-up. Dedicated long-term clinical studies are necessary to fully evaluate the endocrine consequences of preoperative ERFA. Whether such conditioning could modify the need for complex anastomotic reconstruction remains a hypothesis that requires dedicated clinical investigation.

One limitation of this study is that it did not include other types of glue in the comparison, such as neoprene-based glue, which has been reported as a better sealant compared to other alternatives in recent studies led by Dr. Mazzaferro ^12^. However, the cyanoacrylate glue used in this study remains the most current used technique. Despite this limitation, these reported outcomes of ERFA are solid on their own, showing a significant degree of atrophy and reduction in the risk of fistula.

Methodologically, the study is strengthened by its randomized design, use of a large-animal model with anatomy and physiology closely resembling humans, standardized surgical procedures supported by multimedia documentation, and blinded histopathological assessment. The outcomes were assessed comprehensively, including early histology, necropsy findings, biochemical markers, and clinical surrogates, providing robust evidence of the biological and safety profile of EFRA. The limitations of this study include its relatively small sample size, absence of a true pancreato-enteric anastomosis, short follow-up for long-term endocrine outcomes, and lack of comparison against other types of glue. Nevertheless, the consistent early histological effect and reduced macroscopic complications support the clinical relevance of the primary prophylactic mechanism of ERFA.

Future research should aim to optimize the parameters of ERFA, evaluate long-term endocrine and exocrine function, and assess clinical efficacy in high-risk patients undergoing pancreatoduodenectomy. Comparative studies with other energy-based modalities may also refine technique selection. The concept of preoperative pancreatic conditioning could also be applied in neoadjuvant or preoperative planning pathways; such strategies could potentially reduce the incidence of POPF while preserving endocrine function.

Aside from its immediate implications in preventing POPF, the biological remodeling induced by preoperative ductal ablation may have broader translational relevance. Experimental studies in KRAS-driven murine models, including the work of Cáceres et al. ^31^, have shown that complete MPD ligation leads to profound apoptosis-mediated acinar atrophy and a marked reduction in high-grade PanINs and other preneoplastic lesions in the obstructed segment. Sustained depletion of the acinar compartment can modify the biological substrate that supports KRAS-driven neoplastic progression. Considering that EFRA can achieve controlled and homogeneous exocrine involution, future research should explore whether preoperative pancreatic conditioning could also influence long-term oncologic outcomes in high-risk or genetically predisposed settings aside from reducing the fistula risk. Such investigations would expand the conceptual framework of ERFA beyond perioperative morbidity mitigation and open new avenues in the biological modulation of the pancreatic remnant.

This preclinical study demonstrates that preoperative MPD occlusion via EFRA is feasible, safe, and capable of inducing stable exocrine pancreatic atrophy prior to pancreatic transection. Compared to cyanoacrylate-based duct occlusion, ERFA produced more consistent and controlled early pancreatic remodeling, resulting in fewer macroscopic complications associated with fistula formation. These findings support the concept of preoperative pancreatic conditioning as a biologically oriented strategy for POPF risk modulation. Further translational and clinical studies are necessary to determine its safety, indications, potential integration with current surgical practice, and influence in oncologic settings.

## Data Availability

All data produced in the present study are available upon reasonable request to the authors

## Acknowledgments

The authors would like to thank the CMCiB facility center for their support.

## Provenance and peer review

Not commissioned, externally peer-reviewed

